# Effects of a social network modification of a community-based family planning intervention on contraceptive use among adolescent wives in Niger: a cluster randomized controlled trial

**DOI:** 10.64898/2026.04.29.26352112

**Authors:** Holly Baker, Shweta Tomar, Amani Hachimou, Kadidiatou Boubacar Moussa, Jennifer Gayles, Rebecka Lundgren

## Abstract

Niger has the world’s highest adolescent fertility rate. Social network (SN) approaches to family planning may improve intervention impact through diffusion beyond direct beneficiaries. We tested a social network modification of a community-based family planning intervention to increase contraceptive use compared to standard implementation and control.Three-arm cluster-randomized trial in 56 rural villages in Maradi, Niger. Eligible participants were adolescent wives (AW) aged 15–19 with 0–1 children and their husbands. Villages were randomized using covariate-constrained randomization (Minirand): standard Kulawa (100% coverage), SN modification (50% coverage pairing AW-mother-in-law dyads with adopt-a-friend diffusion), or control. Interventions were delivered over 12 months. Blinding of participants and implementers was infeasible; analysts were blinded. Primary outcome was current contraceptive use assessed at endline and analyzed using intention-to-treat difference-in-differences logistic regression adjusting for clustering; no missing data were imputed. ClinicalTrials.gov NCT05777473; trial closed to enrollment.From May 1 to September 30, 2022, 1,538 female AW were enrolled (517 control, 532 Kulawa, 489 Kulawa SN); 1,396 (90.8%) completed endline (May–August 2024). Compared to control, the SN arm significantly increased contraceptive use (AOR 2.36, 95% CI 1.27–4.44); the standard arm showed no significant effect (AOR 1.36, 95% CI 0.76–2.41). Within SN villages, both non-participants (AOR 2.66, 95% CI 1.25–5.70) and direct participants (AOR 2.10, 95% CI 0.99–4.44) showed increased use versus control, demonstrating behavioral diffusion. No intervention-related adverse events were observed in any arm. An SN approach targeting AWs, husbands, mothers-in-law, and adopted friends achieved greater effects than standard implementation despite 50% lower coverage, with evidence of diffusion to non-participants. Leveraging social networks may improve impact of family planning programs in high-fertility settings.

## Introduction

The reproductive health challenges faced by Niger are some of the most serious in the world. Niger has the world’s highest rate of child marriage, with 76% of women aged 20–24 married before age 18, and the highest adolescent fertility rate at 186 births per 1,000 adolescents aged 15–19(1, 2). Modern contraceptive prevalence remains critically low. Only 11% among married women nationally and approximately 6% among married adolescents(3-5) report using contraception. While structural barriers, including limited healthcare access, contribute to these patterns, research consistently identifies social norms as the predominant factor sustaining high fertility: pronatalist expectations, family pressure for early childbearing, and stigma surrounding contraceptive use create powerful disincentives for adoption of contraception even when methods are available(6, 7)

Given the importance of social normative factors, efforts to increase family planning uptake in contexts such as Niger have increasingly turned to social network approaches(8, 9). Social network and social norms theory distinguishes between two mechanisms: descriptive norms and injunctive norms(10-12). Descriptive norms refer to perceptions of what others do, or the observed prevalence of a behavior within one’s reference group. Injunctive norms refer to perceptions of what others approve or disapprove of, or the social expectations and sanctions that govern behavior. Descriptive norms operate through social learning: as individuals observe others in their community successfully adopting family planning, they gain information about feasibility, safety, and social acceptability, increasing their own likelihood of adoption. Injunctive norms, by contrast, operate through social influence, and reflect perceived social expectations and sanctions(11, 12); in high-fertility settings, these often function as barriers, manifesting as stigma around contraceptive use, pressure to demonstrate fertility, and social penalties for deviating from pronatalist expectations(6, 7). Multilevel analyses across sub-Saharan Africa confirm that both mechanisms independently predict contraceptive use beyond individual attitudes(13).

Existing evidence suggests that both descriptive and injunctive norms create potential for substantial behavioral diffusion through social networks. In Nepal, women indirectly exposed to a national family planning mass media campaign through social contacts reported significant attitude and behavior changes(14). Across sub-Saharan Africa, quasi-experimental analysis of Demographic and Health Survey data from multiple countries found that women exposed to family planning messages through interpersonal networks reported approximately 6–7 percentage point increases in modern contraceptive use, with effects operating through a talk-to-use pathway in which information from social contacts translated into behavioral adoption(15). Such diffusion occurs as intervention participants share information, leveraging descriptive norms, and as shifting community practices gradually reduce the social costs of adoption by weakening restrictive injunctive norms.

Complex contagion theory provides further guidance for intervention design(16). With simple contagion, information is readily transmitted through any informal connection. For behaviors requiring social reinforcement, including contraceptive adoption in normatively restrictive settings, information spreads most effectively through closely connected, overlapping network ties where individuals receive validation from multiple trusted sources(16-18). Field experiments demonstrate that seeding behavior change through connected pairs rather than isolated individuals accelerates adoption and social reinforcement(19).

For adolescent wives (AW) in patrilocal settings like Niger, reproductive decisions occur within hierarchical household structures where mothers-in-law often exercise substantial authority over daughters-in-law’s mobility, healthcare access, and fertility timing(20, 21). Recent qualitative research from Uganda confirms that social networks shape contraceptive decision-making through complex pathways involving both supportive and constraining ties(22). Engaging influential network members, including mothers-in-law, may therefore accelerate normative change while reducing interpersonal barriers that young wives face when seeking services independently(23, 24).

The Reaching Married Adolescents (RMA) program, developed by Pathfinder International and evaluated in Dosso Region, Niger, provides a foundation for testing social network approaches(25, 26). RMA is a gender-synchronized intervention designed to increase modern contraceptive use and reduce intimate partner violence (IPV) among married adolescent couples. Based on the Theory of Reasoned Action, the intervention includes three components facilitated by community health workers, known as relais communautaires: gender matched monthly household visits, providing information on healthy timing and spacing of pregnancies and contraceptive access; gender-segregated small group discussions addressing reproductive health, gender norms, and couple communication; and community dialogues engaging gatekeepers including religious leaders, parents, and in-laws to foster an environment supportive of contraceptive use. A cluster-randomized trial of the RMA intervention, upon which this study was based, demonstrated that the combined household visit and small group approach significantly increased modern contraceptive use among married adolescent girls(26).

Building on this evidence, we tested a social network modification of RMA within the context of a USAID-funded health project in Maradi Region, Niger, in which RMA was one intervention component. The modification reduced population coverage from 100% to 50% of eligible AW and their husbands while adding mothers-in-law as direct intervention participants, and incorporating an “adopt-a-friend” organized diffusion component(27). This approach hypothesized that targeting relational dyads embedded in closely connected household networks would generate diffusion effects achieving population-level impact despite reduced direct reach.

## Methods

### Study Design and Setting

We conducted a three-arm, parallel, cluster-randomized controlled trial in Maradi Region, Niger. The trial was embedded within Kulawa, a large multi-sector USAID-funded program (2020–2025) implemented by Save the Children, which included a component to increase use of quality family planning services among women of reproductive age across 15 districts in three regions of Niger. The Kulawa family planning programming drew on the RMA approach previously evaluated in Dosso Region. Maradi was selected from three Kulawa project regions based on safety and accessibility; within Maradi, Aguie and Gazaoua districts were selected based on village counts, population demographics, and absence of other family planning programming that could contaminate results. We compared a social network modification of RMA (Kulawa SN) with standard implementation (Kulawa) and an untreated control (ClinicalTrials.gov NCT05777473). The study was approved by the ethics review boards of the University of California San Diego (protocol number 201921) and the Niger Ministry of Health.

### Participants

Villages were eligible if they were rural, Hausa- or Zarma-speaking, had a population size consistent with 20–80 married low-parity adolescents, and were located within 5 km of a health facility. From approximately 200 eligible villages, 51 were randomly selected and allocated to study arms. AW were eligible if married, aged 15–19 years, had 0–1 children, and had a husband available in the village to participate along with her. Although participants aged 15–17 were minors, parental consent was not required as married adolescents are recognized as emancipated under Nigerien law. Verbal informed consent was obtained from all participants following World Health Organization guidelines for research on violence against women.

### Randomization and Masking

Villages were randomized by our university team 1:1:1 to study arms using the Minirand package in R, stratified by population size, number of AW, and presence of a school. Nine villages with fewer than 10 eligible participants were replaced, and five additional villages were added to achieve target sample size, yielding 21 Kulawa villages, 18 Kulawa SN villages, and 17 control villages. Analysts were blinded to allocation until primary analyses were complete.

### Interventions

Both interventions were delivered by *relais communautaires* and directors of integrated health centers, with content adapted from the previously evaluated RMA program. Implementation followed a quarterly cycle over 12 months beginning February 2023. SA Figure A provides a breakdown of intervention activities.

**Standard Kulawa** was a condensed version of the previously evaluated RMA intervention, endorsed by the Niger Ministry of Health for delivery through existing health structures. It engaged all eligible AW (maximum 50 per village) and their husbands through three components delivered on a quarterly cycle over 12 months, ensuring monthly contact through rotating activities. Home visits conducted separately for both AW and their husbands were conducted by gender-matched relais communautaires in participants’ homes, lasting 45–60 minutes per session and using illustrated, story-based tools to deliver education on pregnancy, healthy timing and spacing of pregnancies, modern contraceptive methods, and referrals to health facilities. Gender-segregated small group discussions for both AW and their husbands (7–8 same-sex peers) facilitated by same-sex relais communautaires fostered social cohesion, built trust, and promoted reflective dialogue on contraception and reproductive health. Community dialogues, facilitated by the director of the nearest integrated health center, engaged all community members in discussion and reflection on family planning, gender roles, and couple communication; village chiefs and religious leaders encouraged attendance.

**Kulawa SN** modified the standard approach in three ways. First, only 50% of eligible AW (maximum 25 per village) and their husbands were randomly selected to receive the intervention, with the remaining 50% serving as within-village non-participants for assessing diffusion effects.

Second, each selected AW was paired with her mother-in-law, who became a direct intervention participant. Mothers-in-law participated in select, non-sensitive home visits with daughters-in-law. They then attended their own separate small group discussions with discussion guides adapted from those for AW to integrate self-reflection on their own experiences as young mothers, their current role and influence within the family, and the benefits of healthy birth spacing for their daughters-in-law, grandchildren, and themselves. Formative research, including 24 qualitative social network interviews and 10 focus group discussions in proximal villages, indicated that mothers-in-law exercise the most authority over married adolescents’ lives and reproductive decisions in this patrilocal context. Mothers-in-law influence norms and behaviors through multiple pathways: their own social networks, their authority over sons and daughters-in-law, and their influence over other family members. Engaging them directly aimed to leverage these multiple pathways of influence.

Third, the Kulawa SN arm incorporated an “adopt-a-friend” organized diffusion component integrated into both home visits and small group discussions. At the beginning of the intervention, each participating AW and husband identified one “friend”—a peer not enrolled in the intervention—to share information with throughout the program. Selection criteria were developed using language reflecting social network questions from the formative research to ensure participants identified a trusted contact. At the beginning of each session, *relais communautaires* asked participants whether they had shared information from the previous session with their adopted friend, noting responses as part of routine monitoring, and ended each session with a reminder to discuss new information with their friend. This structured mechanism aimed to systematize the sharing of information and ideas through existing peer networks, extending intervention reach beyond direct participants through organized rather than solely passive diffusion.

**Control** villages received no intervention during the study period. A comprehensive description of the study design, including details on the intervention are available online(28).

### Data Collection

Baseline surveys were conducted May–September 2022; 1,538 AW completed interviews. Endline assessments occurred May–August 2024, after the completion of one cycle (12 months) of intervention implementation, with 1,396 respondents retained (90.8%). The study surveyed all eligible AW within each village, regardless of their participation status in the intervention. Survey instruments drew on validated measures from Demographic and Health Surveys, were translated from English to French, back-translated for quality assurance, pilot tested, and then administered verbally in Hausa by trained research assistants. Interviews lasted 60–80 minutes and were conducted in private locations. Research assistants received seven-day training covering research ethics, informed consent, strategies for addressing participant distress, and data management. See Figure 1 for the trial profile.

**Figure 1.**
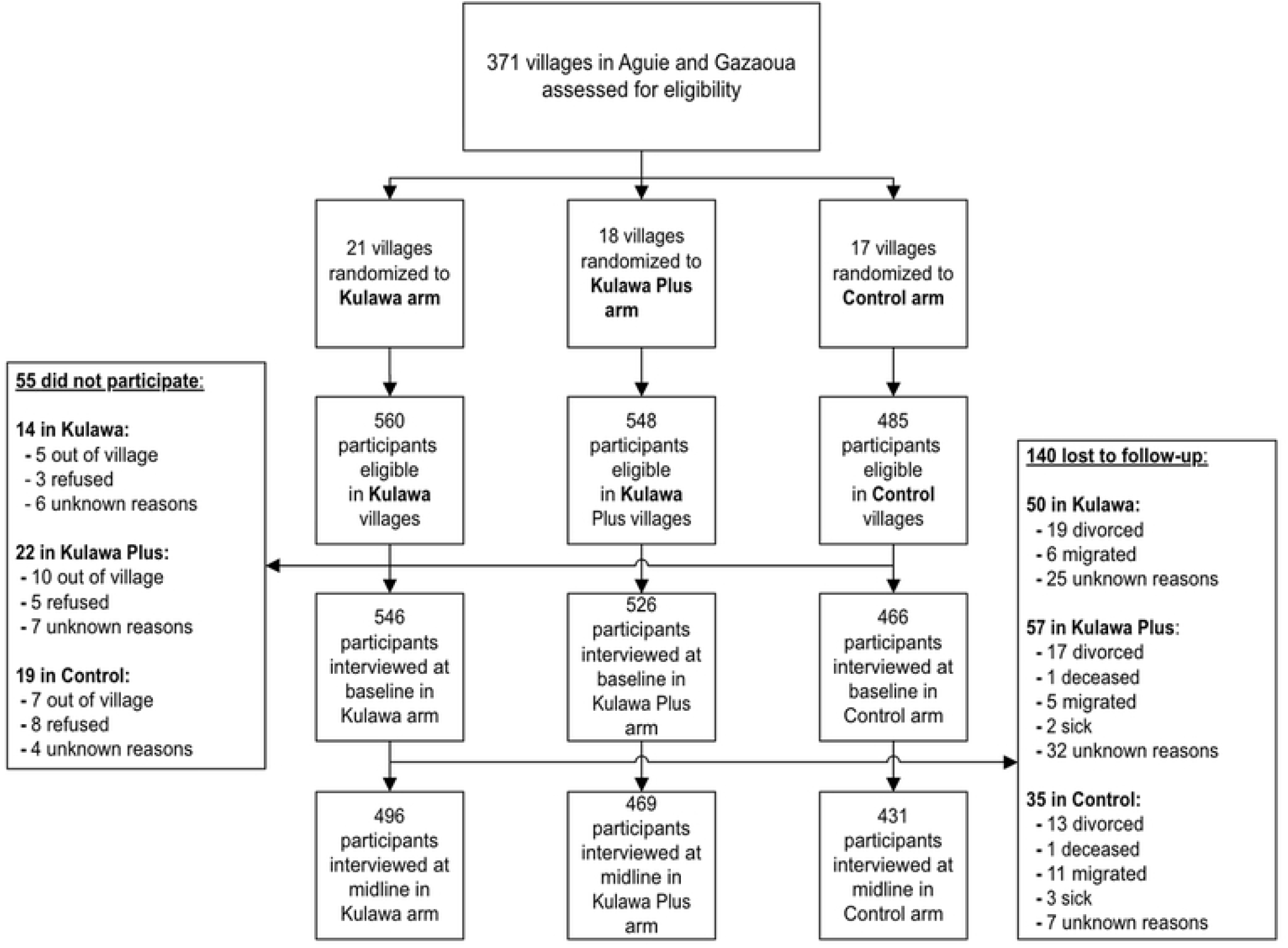
Trial Profile.

### Measures

**Current contraceptive use** (primary outcome) was assessed through two sequential questions. Respondents were first asked: “Have you or your husband ever done something or used any method to space or delay getting pregnant?” Those responding affirmatively were then asked: “Are you or your husband currently doing something or using any method to space or delay getting pregnant?” Affirmative response to the second question indicated current use; negative response to either question indicated non-use. Given the study’s focus on behaviors to space or delay pregnancy in a context where method mix is limited and traditional methods are common, we included any form of contraception rather than restricting to modern methods. Sensitivity analyses with only modern methods were conducted separately and are shown in the online appendix. Respondents who were pregnant at the time of interview or who declined to answer were categorized separately and handled through sensitivity analyses.

**Contraceptive use in past 12 months** (secondary outcome). Respondents who reported ever-use but not current use were asked: “Did you or your husband do something or use any method to space or delay getting pregnant in the past 12 months?” Participants who responded “Yes” to either the current use question or the past 12 months question were considered to have used FP in the past 12 months. This variable captured both ongoing contraceptive use and any use that occurred within the 12-month period but may have since been discontinued.

**Intention to use contraception in next 12 months** (secondary outcome) was assessed by asking the respondents: “Will you continue to use the current FP method over the next 12 months to avoid or delay pregnancy?” or “Will you use a family planning method in the next 12 months to avoid or delay pregnancy?” depending on their current use status. A “Yes” response to either of these questions indicated that the AW intends to use an FP method in the next 12 months. This measure captured cognitive readiness for adoption among current non-users and sustained commitment among current users.

### Sample Size

Based on RMA trial data, we assumed 11% baseline contraceptive prevalence and an effect size of a 24% increase in contraceptive use in the standard Kulawa (Arm 1) vs the control (Arm 3) group, and similar increase in the Kulawa SN (Arm 2). We powered the study to detect diffusion effects among non-participants in the Kulawa SN arm. Assuming non-participants would achieve 50% of the participant effect (12 percentage point increase from 11% to 23%), with ICC=0.05, 90% confidence, and 80% power, we required 225 non-participants per arm. With 50% coverage in Kulawa SN, this required 450 AW per arm; assuming 10% non-response, we targeted 510 per arm (1,530 total).

### Statistical Analysis

Analyses followed a pre-specified plan using difference-in-differences with multilevel mixed-effects logistic regression accounting for clustering within villages. Models included time (baseline/endline), treatment arm, time-by-treatment interaction, and covariates associated with loss to follow-up (see Covariates section in appendix). The primary analysis compared Kulawa and Kulawa SN (all eligible AW) against control. Secondary analyses separated Kulawa SN into participants and non-participants to assess diffusion. An intent-to-treat approach was used; no missing data were imputed. Sensitivity analyses examined alternative approaches to handling pregnancy status and declined responses. All analyses used R.

## Results

From approximately 200 villages assessed for eligibility in Aguie and Gazaoua districts, 56 met inclusion criteria and were randomized to study arms. Between May and September 2022, 1,538 AWs were enrolled: 517 in control villages (17 villages), 532 in Kulawa villages (21 villages), and 489 in Kulawa SN villages (18 villages). Within Kulawa SN villages, 245 were non-participants and 244 were randomly selected as direct participants along with their mothers-in-law. At endline (May–August 2024), 1,396 AWs (90.8%) completed follow-up: 431 in control, 496 in Kulawa, and 469 in Kulawa SN villages (233 non-participants, 236 participants).

Table 1 presents baseline characteristics. AWs were similar across study arms, with mean age approximately 17.4 years, mean age at marriage 15.3 years, and mean parity 0.6 children. Husbands were approximately 10 years older (mean 27–29 years). The sample was predominantly Hausa with a Fula minority. Educational attainment was limited, with the majority having attended only Quranic school or no school at all. Most AWs were in monogamous marriages and were not employed for pay. Approximately half of AWs reported household income insufficient to meet basic needs.

**Table 1.** Baseline characteristics of adolescent wives by study arm.

|  | **Control** | **Kulawa** | **Kulawa SN** | **Non-**<br>**participants** | **Participants** |
| --- | --- | --- | --- | --- | --- |
|  | *N* = 431 | *N* = 496 | *N* = 469 | *N* = 233 | *N* = 236 |
|  | Mean (SD) | Mean (SD) | Mean (SD) | Mean (SD) | Mean (SD) |
| Adolescent wife's age | 17.32 (1.4) | 17.54 (1.32) | 17.42 (1.37) | 17.51 (1.35) | 17.34 (1.39) |
| Husband's age | 26.73 (6.88) | 28.64 (7.67) | 26.87 (6.61) | 26.85 (6.28) | 26.89 (6.92) |
| Age at marriage | 15.2 (1.27) | 15.49 (1.21) | 15.25 (1.27) | 15.27 (1.25) | 15.22 (1.28) |
| Parity | 0.62 (0.75) | 0.62 (0.72) | 0.57 (0.70) | 0.60 (0.72) | 0.53 (0.68) |
| Household asset score | 1.71 (1.05) | 1.85 (1.15) | 1.72 (1.00) | 1.63 (1.02) | 1.81 (0.97) |
|  | % (n) | % (n) | % (n) | % (n) | % (n) |
| **Ethnicity** |  |  |  |  |  |
| Hausa | 71.7% (309) | 90.1% (447) | 79.1% (371) | 77.7% (181) | 80.5% (190) |
| Fula | 27.4% (118) | 7.7% (38) | 20.7% (97) | 21.9% (51) | 19.5% (46) |
| **Education** |  |  |  |  |  |
| None | 50.1% (216) | 36.5% (181) | 48.2% (226) | 48.5% (113) | 47.9% (113) |
| Quranic only | 16.0% (69) | 19.8% (98) | 17.5% (82) | 18.0% (42) | 17.0% (40) |
| Modern only | 18.6% (80) | 17.1% (85) | 19.0% (89) | 16.7% (39) | 21.2% (50) |
| Both | 15.3% (66) | 26.6% (132) | 15.4% (72) | 16.7% (39) | 14.0% (33) |
| Monogamous marriage | 91.0% (392) | 87.9% (436) | 87.4% (410) | 86.7% (202) | 88.1% (208) |
| **Income sufficiency** |  |  |  |  |  |
| Sufficient with savings | 3.3% (14) | 10.3% (51) | 14.7% (69) | 15.9% (37) | 13.6% (32) |
| Sufficient, no major difficulties | 48.7% (210) | 37.5% (186) | 47.6% (223) | 47.2% (110) | 47.9% (113) |
| Insufficient with difficulties | 43.4% (187) | 46.2% (229) | 31.8% (149) | 29.6% (69) | 33.9% (80) |
| Insufficient, major difficulties | 4.6% (20) | 6.1% (30) | 6.0% (28) | 7.3% (17) | 4.7% (11) |
| Husband migrated >3 months | 30.9% (133) | 25.4% (126) | 36.5% (171) | 36.9% (86) | 36.0% (85) |

Data are mean (SD) or % (n). Kulawa SN non-participants and participants shown separately within Kulawa SN overall.

At baseline, current contraceptive use was low across arms: 11.4% in control, 14.7% in Kulawa, and 10.2% in Kulawa SN villages (Table 2). Of those who reported contraceptive use at baseline, 68% used modern methods and 32% used traditional methods (SA Table A). A subset of respondents was pregnant at baseline (11–15% across arms) or declined to answer contraceptive use questions (3–6%). Intention to use contraception in the next 12 months was approximately 20% across all arms.

**Table 2.** Outcome distributions at baseline and endline by study arm.

|  | Baseline |  |  |  |  | Endline |  |  |  |  |
| --- | --- | --- | --- | --- | --- | --- | --- | --- | --- | --- |
|  | Control | Kulawa | KSN | Non-P | Part | Control | Kulawa | KSN | Non-P | Part |
|  | % (n) | % (n) | % (n) | % (n) | % (n) | % (n) | % (n) | % (n) | % (n) | % (n) |
| **Current FP use** |  |  |  |  |  |  |  |  |  |  |
| No | 73.1 (315) | 65.5 (325) | 71.4 (335) | 73.4 (171) | 69.5 (164) | 61.0 (263) | 44.8 (222) | 48.4 (227) | 52.4 (122) | 44.5 (105) |
| Yes | 11.4 (49) | 14.7 (73) | 10.2 (48) | 9.4 (22) | 11.0 (26) | 28.5 (123) | 44.2 (219) | 40.9 (192) | 39.1 (91) | 42.8 (101) |
| Pregnant | 13.0 (56) | 15.3 (76) | 12.8 (60) | 11.2 (26) | 14.4 (34) | 9.7 (42) | 10.5 (52) | 10.0 (47) | 8.6 (20) | 11.4 (27) |
| Declined | 2.6 (11) | 4.4 (22) | 5.5 (26) | 6.0 (14) | 5.1 (12) | 0.7 (3) | 0.6 (3) | 0.6 (3) | 0.0 (0) | 1.3 (3) |
| **Intention to use FP** |  |  |  |  |  |  |  |  |  |  |
| No | 65.4 (282) | 62.9 (312) | 67.2 (315) | 65.7 (153) | 68.6 (162) | 58.0 (250) | 38.7 (192) | 45.2 (212) | 51.1 (119) | 39.4 (93) |
| Yes | 19.5 (84) | 19.8 (98) | 19.8 (93) | 20.6 (48) | 19.1 (45) | 32.0 (138) | 47.2 (234) | 42.9 (201) | 38.2 (89) | 47.5 (112) |
| Don't know | 9.3 (40) | 10.1 (50) | 5.5 (26) | 5.6 (13) | 5.5 (13) | 6.3 (27) | 8.5 (42) | 5.3 (25) | 6.0 (14) | 4.7 (11) |
| Declined | 5.8 (25) | 7.3 (36) | 7.5 (35) | 8.2 (19) | 6.8 (16) | 3.7 (16) | 5.7 (28) | 6.6 (31) | 4.7 (11) | 8.5 (20) |
| **Past 12-month FP use** |  |  |  |  |  |  |  |  |  |  |
| No | 82.4 (355) | 77.2 (383) | 81.7 (383) | 80.7 (188) | 82.6 (195) | 62.7 (270) | 45.2 (224) | 48.8 (229) | 54.5 (127) | 43.2 (102) |
| Yes | 12.1 (52) | 15.7 (78) | 11.1 (52) | 11.2 (26) | 11.0 (26) | 35.5 (153) | 52.2 (259) | 47.8 (224) | 43.8 (102) | 51.7 (122) |
| Declined | 5.6 (24) | 7.1 (35) | 7.3 (34) | 8.2 (19) | 6.4 (15) | 1.9 (8) | 2.6 (13) | 3.4 (16) | 1.7 (4) | 5.1 (12) |

Data are % (n). KSN = Kulawa SN; Non-P = non-participants; Part = participants.

The Kulawa SN intervention significantly increased current contraceptive use compared to control (Table 3). In difference-in-differences analysis excluding respondents pregnant or declining to answer at either time point (n=1,005), AWs in Kulawa SN villages had 2.36 times the odds of current contraceptive use relative to control (95% CI 1.27–4.44, p<0.01). Standard Kulawa showed no significant effect (AOR 1.36, 95% CI 0.76–2.41, p=0.30).

**Table 3.**
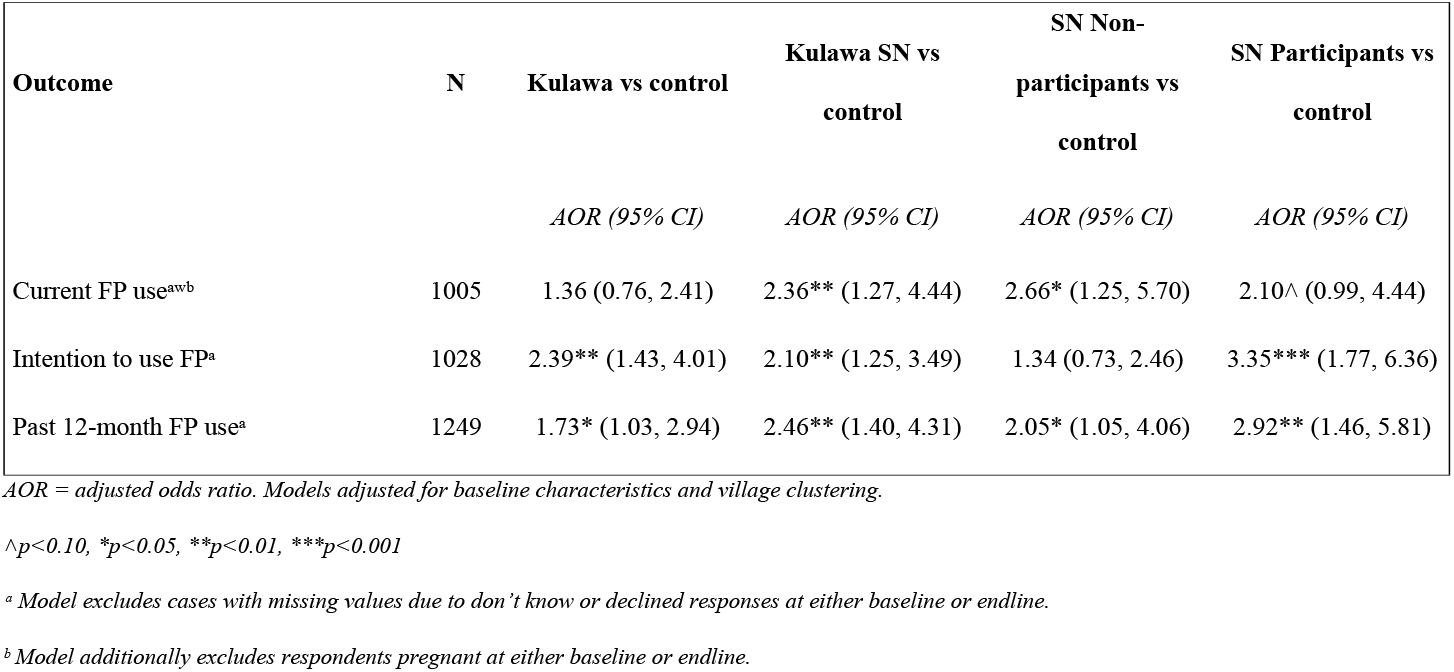
Effect of interventions on contraceptive outcomes: difference-in-differences analysis.

When we separate Kulawa SN participants from non-participants, we find strong evidence of diffusion. Non-participants—AWs residing in Kulawa SN villages who were not randomly selected for intervention— demonstrated odds of contraceptive use compared to control (AOR 2.66, 95% CI 1.25–5.70, p=0.01) at least as high as, if not higher than, direct participants (AOR 2.10, 95% CI 0.99–4.44, p=0.05).

In absolute terms, current contraceptive use at endline reached 28.5% in control villages, 44.2% in Kulawa villages, and 40.9% in Kulawa SN villages (39.1% among non-participants, 42.8% among participants).

Contraceptive use in past 12 months. Among respondents with complete data (n=1,249), Kulawa SN significantly increased odds of past-year contraceptive use compared to control (AOR 2.46, 95% CI 1.40-4.31, p<0.01). Standard Kulawa also showed significant effects (AOR 1.73, 95% CI 1.03–2.94, p=0.04). Within Kulawa SN villages, both non-participants (AOR 2.05, 95% CI 1.05–4.06, p=0.04) and participants (AOR 2.92, 95% CI 1.46– 5.81, p<0.01) showed significantly increased use. At endline, past-year use reached 35.5% in control, 52.2% in Kulawa, and 47.8% in Kulawa SN villages (43.8% non-participants, 51.7% participants). In absolute terms, past-year use increased by approximately 23 percentage points in control compared to 37 percentage points in Kulawa and 37 percentage points in Kulawa SN.

Intention to use contraception in next 12 months. Among respondents with complete data (n=1,028), both interventions significantly increased future contraceptive use intentions. Standard Kulawa more than doubled the odds of intending future use (AOR 2.39, 95% CI 1.43–4.01, p<0.01), as did Kulawa SN overall (AOR 2.10, 95% CI 1.25–3.49, p<0.01). Diffusion patterns differed markedly for intentions compared to behavior: direct participants showed the strongest effect (AOR 3.35, 95% CI 1.77–6.36, p<0.001), while non-participants showed no significant increase (AOR 1.34, 95% CI 0.73–2.46, p=0.35). At endline, intention to use reached 32.0% in control, 47.2% in Kulawa, and 42.9% in Kulawa SN villages (38.2% non-participants, 47.5% participants). This divergence between intention and behavior outcomes is notable: while contraceptive use diffused to non-participants, stated intentions did not, suggesting that diffusion may operate primarily through observational and normative channels that facilitate behavioral adoption without necessarily shifting cognitive commitment to future use.

Results were robust across sensitivity analyses examining alternative approaches to handling pregnancy status and declined responses (Supplementary Table B). For current contraceptive use, effect estimates for Kulawa SN remained significant whether pregnant respondents were excluded (primary model, AOR 2.36), coded as non-users (AOR 2.16, 95% CI 1.27–3.71), or when declined responses were additionally coded as non-users (AOR 2.16, 95% CI 1.27–3.67). The pattern of effects among non-participants persisted across all specifications. For intention to use, coding “don’t know” responses as negative attenuated effect sizes but maintained significance for direct participants (AOR 2.53, 95% CI 1.39–4.62) and overall Kulawa SN effects (AOR 1.70, 95% CI 1.04–2.75). Results were also robust when restricting the outcome to modern contraceptive methods only (Supplementary Table B).

## Discussion

Can family planning programs achieve greater impact by reaching fewer people? This three-arm cluster-randomized trial among AWs in rural Niger suggests they can under the right conditions. A social network modification that engaged only half of eligible AW and their husbands, paired with mothers-in-law, and employing an adopt-a-friend peer diffusion strategy, achieved significantly greater effects on contraceptive use than standard implementation with full coverage. AWs who received no direct intervention showed contraceptive increases comparable to direct participants, providing experimental evidence that behavioral diffusion through closely connected relational networks can extend program reach beyond those directly served.

These findings emerge from one of the most challenging contexts globally for family planning programming. Niger has the world’s highest adolescent fertility rate, high rates of child marriage, and modern contraceptive prevalence of only 6% among married adolescents(1, 2). Pronatalist expectations are deeply embedded in social and religious institutions, and fertility decisions for AWs occur within hierarchical household structures where mothers-in-law exercise considerable authority over daughters-in-law’s mobility, healthcare access, and reproductive timing(20, 21).That a network-targeted approach succeeded in this context—where normative barriers predominate and structural constraints compound individual-level challenges—suggests considerable potential for similar approaches in other high-fertility settings.

The magnitude of effects merits emphasis. Kulawa SN more than doubled the odds of current contraceptive use compared to control (AOR 2.36, 95% CI 1.27-4.44), while standard Kulawa with 100% coverage showed no significant effect (AOR 1.36, 95% CI 0.76-2.41). Within Kulawa SN villages, both non-participants (AOR 2.66, 95% CI 1.25-5.70) and direct participants (AOR 2.10, 95% CI 0.99-4.44) showed increased contraceptive use compared to control, with non-participants showing significant effects despite receiving no direct programming. In absolute terms, current contraceptive use at endline reached 40.9% in Kulawa SN villages versus 28.5% in control, a 12 percentage point difference achieved while serving only half the eligible population.

These patterns extend assumptions about dose-response relationships in behavior change interventions to consideration of who is being identified in intervention strategies. Standard Kulawa, despite engaging 100% of eligible participants, showed no significant effect on current contraceptive use, while Kulawa SN with 50% coverage achieved more than double the odds of use. Two mechanisms may explain this promising finding. First, engaging mothers-in-law as direct participants may have addressed a critical barrier that standard programming leaves intact: household-level gatekeeping of daughters-in-law’s reproductive autonomy(20). When mothers-in-law are included in the intervention, they are not merely informed about family planning but actively participate in the normative dialogue, potentially transforming them from gatekeepers resisting change to advocates facilitating it. Second, concentrating intervention resources on relational dyads embedded in closely connected networks may have generated reinforcing social dynamics—consistent with complex contagion theory(16-18)-that amplified impact beyond what diffuse, universal coverage could achieve.

The evidence for diffusion to non-participants represents a central contribution of this trial. Prior research has documented associations between network characteristics and contraceptive adoption(29, 30), and quasi-experimental studies have estimated diffusion effects in sub-Saharan Africa(15) and Nepal(14). However, these studies could not experimentally isolate diffusion from direct exposure. By randomizing intervention participation within Kulawa SN villages, our design enabled unconfounded estimation of effects among non-participants who received no direct programming. The resulting effect size (AOR 2.66) exceeded our conservative power assumptions and suggests that diffusion potential in cohesive network structures may be substantial when interventions successfully shift normative expectations among influential actors.

The divergent patterns across behavioral and cognitive outcomes illuminate potential mechanisms of diffusion. For contraceptive use, both current and past 12-month, non-participants showed significant increases comparable to participants. For intentions, however, only direct participants showed effects while non-participants did not differ from controls. This pattern is consistent with diffusion operating primarily through descriptive norm channels: as community members observe successful adoption by others, they gain information about feasibility and social acceptability that facilitates their own adoption, even without corresponding shifts in stated intentions(10, 11, 30) The practical implication is that behavior change through social diffusion may not require comprehensive attitudinal change among all community members; reducing the social costs of adoption through visible examples may be sufficient to shift behavior.

These findings also align with complex contagion theory, which posits that complex behaviors requiring social reinforcement spread most effectively through closely connected, overlapping network ties rather than weak bridges.^17–19^ Contraceptive adoption in normatively restrictive settings requires validation from multiple trusted sources to overcome social sanctions(6, 7). By engaging mother-in-law and daughter-in-law pairs, the intervention may have created reinforcing channels within household networks that extended influence to other family members and community contacts. The adopt-a-friend component may have further facilitated this process by providing structured pathways for peer-to-peer information sharing.

The original RMA trial in Dosso Region demonstrated that combined monthly household visits and small group discussions significantly increased contraceptive use among married adolescents(26). In the current trial, Kulawa adapted the RMA approach for scale within routine programming, reducing intervention intensity from monthly to quarterly activities. This reduced dosage may partially explain the null effect of standard Kulawa on current contraceptive use; the intervention as implemented may have been insufficient to overcome normative barriers without additional network-targeting components. The contrast between RMA’s demonstrated efficacy and standard Kulawa’s null effect underscores the well-documented challenges of maintaining intervention fidelity and impact during programmatic scale-up.

A key limitation of our design is that we cannot disentangle the relative contributions of the modifications distinguishing Kulawa SN from standard Kulawa: the addition of mothers-in-law as direct participants and the adopt-a-friend organized diffusion component. Both may have contributed to observed effects—mothers-in-law through their gatekeeping authority over daughters-in-law’s reproductive decisions(20), and adopt-a-friend through explicit facilitation of social learning among peers. Factorial designs would be required to isolate these mechanisms. Additionally, the 50% coverage in Kulawa SN may itself have concentrated intervention resources and fostered stronger group cohesion compared to universal coverage.

Several additional limitations warrant consideration. Contraceptive use was assessed by self-report, which may be subject to social desirability bias; however, any such bias would likely operate similarly across randomized arms. The 12-month follow-up captures relatively short-term effects; whether diffusion-driven behavior change is sustained requires longer observation. Pregnancy and declined responses necessitated analytic decisions about handling missing data; sensitivity analyses examining alternative specifications yielded consistent findings. The trial was conducted in two districts of Maradi Region among a predominantly Hausa population; generalizability to other ethnic groups, urban settings, or regions with different kinship structures requires further investigation. The noteworthy increase in contraceptive use in the control arm (from 11.4% to 28.5%) may reflect secular trends, broader Kulawa programming effects in the region, or temporal changes in contraceptive access, but does not compromise between-arm comparisons given the randomized design. Finally, we could not directly measure hypothesized diffusion mechanisms including information sharing between participants and non-participants or normative change among mothers-in-law; secondary analyses of social network data will attempt to address these questions in subsequent publications.

These findings carry implications for family planning programming in high-fertility, normatively constrained settings. Engaging influential household members, particularly mothers-in-law in patrilocal contexts, as direct intervention participants rather than peripheral targets may address gatekeeping barriers that limit program effectiveness. Strategic targeting of relational clusters can generate diffusion effects that extend reach beyond direct participants, potentially improving both impact and cost-effectiveness. The divergence between behavioral and cognitive outcomes suggests that facilitating adoption through normative diffusion may not require comprehensive attitudinal shifts: visible behavior change among respected community members may be sufficient to lower social barriers for others.

In conclusion, a social network modification of a community-based family planning intervention achieved greater effects on contraceptive use than standard implementation despite engaging only half the eligible population. Evidence of behavioral diffusion to non-participants supports the potential of network-targeted approaches to amplify impact while reducing implementation burden in settings where normative barriers predominate. Further research should examine durability of effects, cost-effectiveness, the relative contributions of mother-in-law engagement and peer diffusion mechanisms, and applicability to other health behaviors requiring social reinforcement.

## Data Availability

Data Sharing De-identified individual participant data will be made available upon reasonable request to the corresponding author, subject to a data use agreement, beginning 12 months after publication

## Acknowledgments

We thank the adolescent wives and their families who participated in this study, Save the Children Niger for their hard work on the implementation, GRADE Africa for their tireless work on the survey development and data collection, the community health workers and health center staff who delivered the interventions, and the research assistants who collected data. We acknowledge the Niger Ministry of Health for their support of this research.

